# Impact of breastfeeding on stunting : A systematic review and meta-analysis of observational studies in Ethiopia

**DOI:** 10.1101/2025.09.02.25334897

**Authors:** Fillmon Kidus Ebuy, Tsegaye Getachw Mekonen, Tesema Kecha Meskele

## Abstract

**Introduction:** Not providing breastfeeding reduce key nutrient that support child’s optimal growth and immune boosting proteins component helps prevent a child from various disease, and consequently increases the risk of stunting. Studies showed inconsistency regarding breastfeeding statistical significant association with child stunting.

**Objective:** The initial aim of the systematic review and meta analysis was to synthesis the pooled effect size of researches on the impact of breastfeeding on stunting and the subsequent aim of the study was to evaluate the consistency of statistical significance.

**Method:** An International Prospective Register of Systemic Review received the prepared review protocol and registered it. The Preferred Reporting Items for Systemic Review and Meta-Analysis guidelines were followed in the direction of this study. Two reviewers independently carried out the data extraction. An electronic bibliographic search was conducted using Google Scholar, PUB Med, MEDLINE, EMBASE, LILACS, and the WHO journal. Using the Risk of Bias in Non-Randomized Studies of Intervention tool, the study examines risk of bias.To evaluate the aggregate effect, the study used a random effect meta analysis approach. Cochran’s Q and I^2^ statistics were used to evaluate statistical heterogeneity. In order to determine the cause of heterogeneity, subgroup analysis was conducted.

Begg’s funnel plot and Eggers regression test were used to assess publication bias; the leave-one-study approach was used to conduct sensitivity analysis; and the Grading of Recommendation Assessment, Development, and Evaluation framework was used to assess certainty evidence.

**Result:** Based one litrature search we included nine studies based on established criteria. The overall pooled effect measure demonstrated a statistically significant association [OR = 1.54, 95%CI(0.94-2.13), P=<0.00]. Heterogeneity, revealed significant with [Q=34.44, degree of freedom =8 and P < 0.001] while, the I ^2^ statistic test showed [I^2^ = 86%]. The overall studies’ risk of bias was deemed to be moderate. According to subgroup analysis, the source of the heterogeneity for the studies included in the review were study location, study design and sample size. The results of sensitivity analysis were deemed not robust. Publication bias of Begg’s funnel plot reported asymmetry and Egger’s regression test indicated presence of publication bias with [OR= 1.38, 95%CI (0.17-2.59) P= 0.03]. We applied the trim and fill method to adjust and enhance the symmetry. Indicated the adjusted pooled effect of [OR= 1.31, 95%CI (0.79-1.82) P= <0.001]. Certainty evidence of the studies was found to be moderate.

**Conclusion and recommendation:** According to the results of the meta-analysis, not breastfeeding is a risk factor for a higher incidence of stunting in Ethiopia. Additionally, confirmed presence of heterogeneity among the studies. In order to encourage breastfeeding and prevent the high prevalence of stunting in Ethiopia, health officials should incorporate policies into their nutrition programs.

## Introduction

Stunting has immediate effects on mortality, morbidity, and disability, and long-term effects include determinant effects on adult body size, blood vessel and metabolic disease, intellectual capacity, economic production, and reproductive potential. Stunting is defined as a child’s height or length that is too low for their age, as determined by the LfA (Length for Age)-Z score, which is 2 standard deviations (SD) of the WHO (World Health Organization) child growth standards median (1).

About half of all fatalities among children under five worldwide are attributed to under-nutrition, according to the United Nations Children’s Fund (UNICIFE) and the World Health Organization’s best estimates (1). In developing countries, stunting or linear growth retardation in young children is often caused by malnutrition (5). Stunting was more prevalent in Saharan African countries (33%) than in West Africa (31.8%), Southern Africa (30.6%), and Central Africa (28.8%), with a prevalence of 39% higher in East Africa, according to a meta-analysis pooled estimate (6). Ethiopia has the greatest rate of undernutrition among children under five, with 37–42% of the population experiencing stunting (7,8).

Breastfeeding lowers the risk of stunting in children, and studies in LMICs (Lower Middle Income Countries) have demonstrated a dose response association between breast feeding and a lower risk of stunting (9–12). Since breastfeeding is the main way to ensure that children receive adequate nutrients and is the basis for future breastfeeding success, it can lower the risk of stunting (13).

According to some study done in Ethiopia, there were statistically significant correlation between stunting and breastfeeding(15-17, 20-22). Some studies also showed breastfeeding as a not statistical significant association with child stunting(14, 18-19). Other study also show breast feeding as inverse relation with child stunting(22). In Ethiopia, no systematic review and meta-analysis examining the impact of breastfeeding on stunting has been done before. Therefore, it is crucial to undertake a systematic review and meta-analysis of Ethiopian studies in order to produce tangible evidence on the impact of breastfeeding on stunting. Therefore, by examining observational studies carried out in Ethiopia, this systematic review and meta-analysis will ascertain the pooled effect and evaluate the consistency of the data on the effect of breastfeeding on stunting. The study’s conclusions will assist decision-makers and relevant health sector entities in putting in to practice the successful breastfeeding intervention to prevent risk of stunting.

## Method and material

The protocol which was produced and registered in PROSPRO (International Prospective Register of Systematic Review; it is possible to accessed here https://www.crd.york.ac.uk/PROSPERO/view/CRD420251108544). Furthermore, the PRISIMA (Preferred Reporting Items for Systemic Review and Meta-Analysis) guidelines were followed in this systematic review and meta-analysis (23).

### Search strategy

We conducted an electronic bibliographic search in the following data base: PUBMed, MEDLINE, EMBASE, LILACS, Google scholar, and WHO journal data bases. We used search strategy in PUBMed and EMBASE as follow : ((((("breast feeding"[MeSH Terms] AND "growth disorders"[MeSH Terms]) OR "malnutrition"[MeSH Terms]) AND "ethiopia"[MeSH Terms] AND ("loattrfree full text"[Filter] AND ((ffrft[Filter]) AND (2000/1/1:2025/7/1[pdat])) In Google scholar : "breast-feeding""stunting” “under-nutrition” “under-growth” "Ethiopia" “Cohort” “cross-sectional” “case-control” “01/01/2000:2025/07/01”

### Selection process

To choose possible full-text articles for more thorough examination, two impartial reviewers(FK and TG) were first skim titles and abstracts. The complete text of the paper was retrieved through connected data bases and thoroughly examined for inclusion by the two reviewers after title and abstract have been approved. Based on the year of publication and subject regions, studies that are published by the same researchers and examine the same factor was assessed for any duplication data. The study with the biggest data set was used for meta analysis where there were duplicates. Discussions with the third reviewer (TK) were used to settle any disagreements.

### Eligibility criteria

#### Study type

Inclusion criteria : Observational studies design, published starting January, 2000 until July, 2025 and study conducted in Ethiopia Exclusion criteria: Studies only with abstracts and study conducted in other countries

#### Study population

Inclusion : All stunted under five years old childeren

Exclusion : All stunted children beyond the age of five years old Outcome of interest

Inclusion: The primary outcome is stunted under five year children

Exclusion: Stunted children beyond the age five years old

Exposure of interest :

Inclusion: Breastfeeding is the practice of giving breast milk to infants, ages 0 to 59 months, either directly from the mother’s breast or by pumping the milk into a bottle and delivering it to the infants.

Exclusion: feeding of breast milk to children beyond the age of 59 month.

### Data collection process

The included studies were retrieved and calibrated using an excel data extraction tool that the researcher built based on the pilot review. (FK and TG), the two reviewers, separately retrieved data from the eligible research papers in order to assess the study’s quality and synthesize the data. Disagreements were then discussed with TK, the third reviewer, in order to pinpoint and resolve them. The relevant details that were included the following: author, study site, study population, participant demographics, exposure details, study design, outcome, information on risk of bias assessment, sample size, confounding, and effect size measurement. To seek any missing data, the study’s authors were emailed. Contact emails were used to ask study authors for missing data.

### Data Items (Outcome)

The outcome domain and definition is: Stunting is defined as length/height-for-age under – 2 standard deviations (SD) of the WHO child growth standards median referred to as LfA-z-score, meaning that children’s length/height is too low for their age.

### Data Item(Other variable)

Author, country, year of publication, type of study design, sample population, variable and effect of measurement type.

### Study risk of bias assessment

The ROBINS I tool (Risk of Bias in Non-Randomized Studies-of Intervention)(24) was used by two reviewers (FK and TG) to independently assess the risk of bias in included studies. A third reviewer (TK) participated in the discussion to settle any disputes between the two reviewers (FK and TG) regarding the risk of bias in particular studies. Overall, the quality assessment results were used in data synthesis for pre planned sensitivity analysis, to test the effect of removing low-quality studies.

### Synthesis Methods

All meta-analyses were performed using IBM SPSS version 31.0.0. We employed random effect model, because of presence of heterogeneity with in the studies. The summery of effect size for each study was calculated using the Odds ratio (OR) with and P value P< 0.05 as significant value. Statistical heterogeneity was assessed using cochran’s Q statistical test of and I^2^ statistics test to quantify the level of heterogeneity between studies. We considered I^2^ value greater than 50% to be indicative of substantial heterogeneity (25). Sensitivity analysis was conducted using leave - out - one study meta-analysis method to evaluate the influence of each study on the pooled effect size.

#### Subgroup analysis

Due to the number of studies were less than ten we did not performed meta regression analysis. However; we conducted subgroup analysis to identify source of the heterogeneity.

#### Publication bias assessment

We implemented Begg’s modified funnel plot and Egger’s regression test (26) when there are five or more studies evaluating the relationship of a specific factor. Egger’s regression test reported publication bias (P-value < 0.1), we applied the trim and fill method proposed by Duvall and Tweedie to (26) to include missing studies and enhance symmetry. Following the addition of these potentially missing studies, we revised the pooled effect size and its 95% confidence interval. To address gaps in information regarding the methods utilized in the studies included and to retrieve missing statistics, we reached out to authors via the email addresses provided in the publications or access phone directories of their affiliated organizations.

#### Certainty assessment

The assessment of certainty were conducted using the GRADE(Grade of Recommendations Assessment, Development and Evaluation) method(27) and the findings were displayed utilizing GRADEpro software(28). Criteria used to assess the certainty of evidence result, number of study, research design, risk of bias, inconsistencies, indirectness, imprecision, publication bias, confounding, big effect size and dosage response prediction.

## Result

### Study selection

Between June 1, 2025, and July 1, 2025, a literature review was carried out using databases from MEDLINE, PUBMED, LILACS, SCOUPS, Google Scholar, EMBASE, and WHO journal. The review was completed using the PRISMA 2020 reporting system (Preferred Reporting Item for Systemic review and Meta-Analysis). A total of 1258 records pertaining to the review topic were detected, spanning the publication year January, 2000 to July 1, 2025. After eliminating duplicate retrievals of records, 746 records remained, of which 723 were removed during the initial evaluation because their titles were irrelevant. Full text of the remaining 22 records were viewed. Nevertheless, out of the 22 records evaluated for eligibility, 13 studies (29-37, 39-42) were disqualified due to that were deemed irrelevant in light of exposure and outcome. Consequently, nine full-text articles from studies (14-22) were accessed and added to the review (Figure 1). Three case control studies and five cross-sectional studies made up the study design for the review (Table 1).

**Fig 1.**
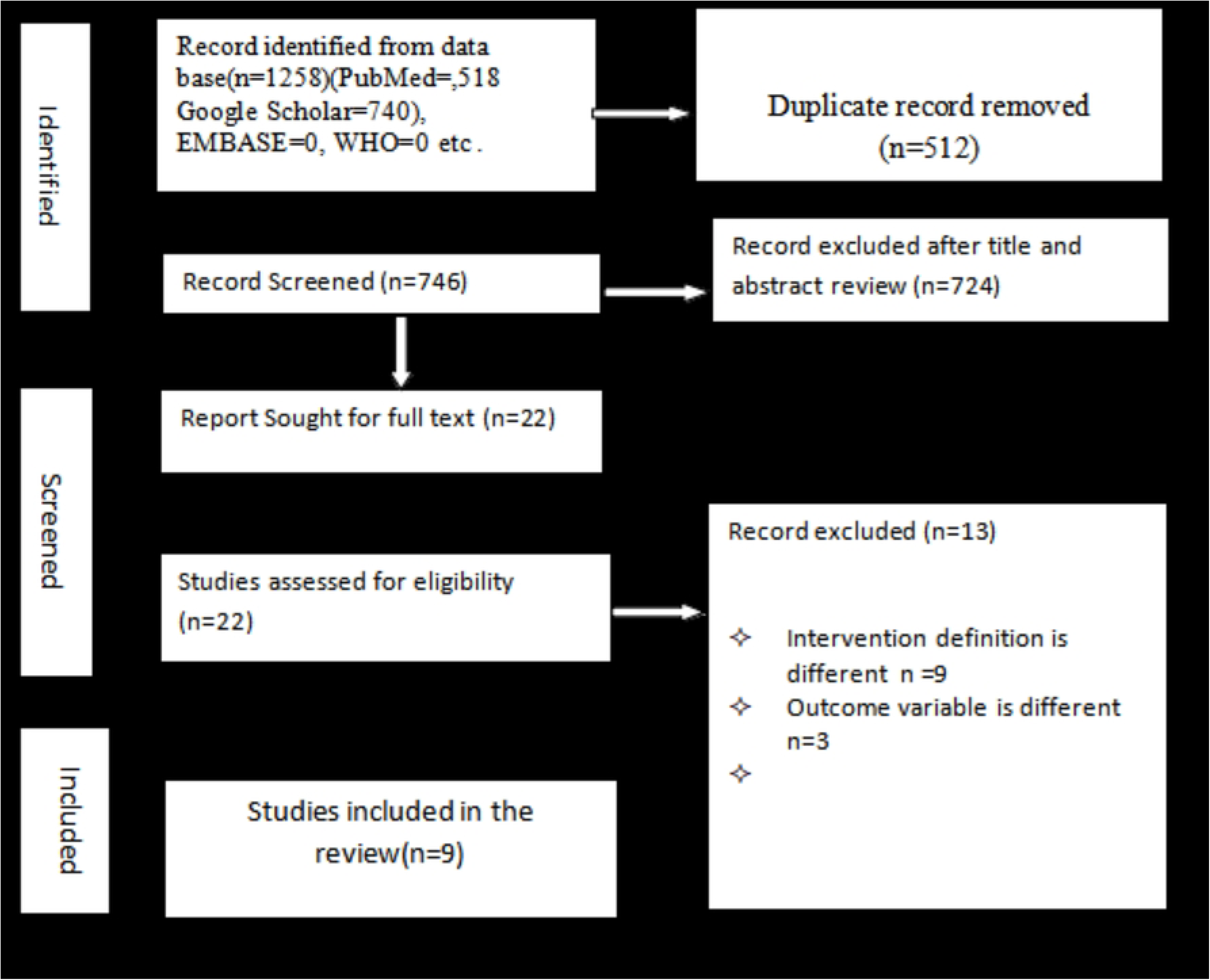
Flow chart of study search results adapted from the PRISMA statement for impact of breastfeeding on stunting : A systematic review and meta-analysis of observational studies in Ethiopia, 2000-2025.

**Table 1.** Characteristics of observational studies included in the systematic review and meta-analysis of impact of breastfeeding on stunting in Ethiopia, 2000-20025.

| Author, Year | Region/location | Study Design | Sample Size | Sampling Method | Effect Measure | Outcome | Intervention |
| --- | --- | --- | --- | --- | --- | --- | --- |
| Kahssay etal[14], 2020 | Afar | Case control | 322 | Simple random | OR | Stunting | Breast Feeding |
| Fikadu etal[15], 2014 | Southern | Case control | 142 | Multistage cluster | OR | Stunting | Breast Feeding |
| Wondmagegne etal[16], 2017 | Oromiya | Case control | 1038 | Simple random | OR | Stunting | Breast Feeding |
| Geberayohanese etal[17], 2022 | Amhara | Cross sectional | 554 | Multistage | OR | Stunting | Breast Feeding |
| Tola etal [18], 2022 | Sidama | Cross sectional | 512 | Multistage cluster | OR | Stunting | Breast Feeding |
| Wolde etal [19], 2014 | Oromiya | Cross sectional | 593 | mullti stage cluster | OR | Stunting | Breast Feeding |
| Bogale etal [20], 2020 | Southern | Cross sectional | 656 | Simple random | OR | Stunting | Breast Feeding |
| Takele etal [21], 2019 | Ethiopia | Cross sectional | 15683 | Multistage cluster | OR | Stunting | Breast Feeding |
| Ahmed etal [22], 2021 | Ethiopia | Cross sectional | 9089 | Multi stage cluster | OR | Stunting | Breast Feeding |

**Table 2.** Subgroup analysis on impact of breastfeeding on stunting : A systematic review and meta-analysis of observational studies in Ethiopia, 2000-2025.

| Subgroup Analysis | Effect size | SE | P- value | Lower CI (95%) | Upper CI 95% | I<sup>2</sup> | Q test(P-value) |
| --- | --- | --- | --- | --- | --- | --- | --- |
| Sampling |  |  |  |  |  |  |  |
| Multi stage cluster | 1.37 | 0.41 | <0.001 | 0.57 | 2.13 |  |  |
| Simple random | 1.78 | 0.29 | <0.001 | 1.27 | 2.36 |  |  |
| Over all subgroup | 1.54 | 0.30 | <0.001 | 0.94 | 2.13 | 86% | <0.001 |
| Location |  |  |  |  |  |  |  |
| Afar | 4.45 | 2.58 | 0.08 | -0.60 | 9.51 |  |  |
| Amhara | 2.25 | 0.75 | <0.001 | 0.77 | 3.72 |  |  |
| Ethiopia | 0.92 | 0.32 | <0.001 | 0.29 | 1.55 |  |  |
| Oromiya | 2.15 | 0.45 | <0.001 | 1.26 | 3.04 |  |  |
| Sidama | 2.86 | 2.54 | 0.26 | -2.12 | 7.84 |  |  |
| Southern | 2.81 | 1.86 | 0.13 | -0.84 | 6.47 |  |  |
| Over all subgroup | 1.54 | 0.30 | <0.001 | 0.94 | 2.13 | 86% | <0.001 |
| Study design |  |  |  |  |  |  |  |
| Case Control | 5.03 | 1.78 | <0.001 | 1.55 | 8.52 |  |  |
| Cross Sectional | 1.42 | 0.29 | <0.001 | 0.86 | 1.98 |  |  |
| Over all subgroup | 1.54 | 0.30 | <0.001 | 0.94 | 2.13 | 86% | <0.001 |
| Sample size |  |  |  |  |  |  |  |
| <1000 | 6.47 | 0.39 | <0.001 | 0.15 | 0.99 |  |  |
| 1000-5000 | 2.14 | 0.47 | <0.001 | 0.21 | 1.12 |  |  |
| >5000 | 0.92 | 0.32 | <0.001 | -1.24 | 0.44 |  |  |
| Over all subgroup | 1.54 | 0.30 | <0.001 | 0.94 | 2.13 | 86% | <0.001 |

### Study Characteristics

The studies were observational in nature, comprising six cross sectional, three case control. Additionally, the studies were conducted in diverse region of Ethiopia (Table 1).

### Risk of bias assessment in studies

The ROBINS I tool, which have seven domains; confounding bias, intervention classification, participant selection, deviation from the intended intervention, risk of bias owing to missing data, outcome measurement, and reported result selection was used to assess the risk of bias in each study. The overall studies’ risk of bias was deemed to be moderate (Figure 2).

**Fig 2.**
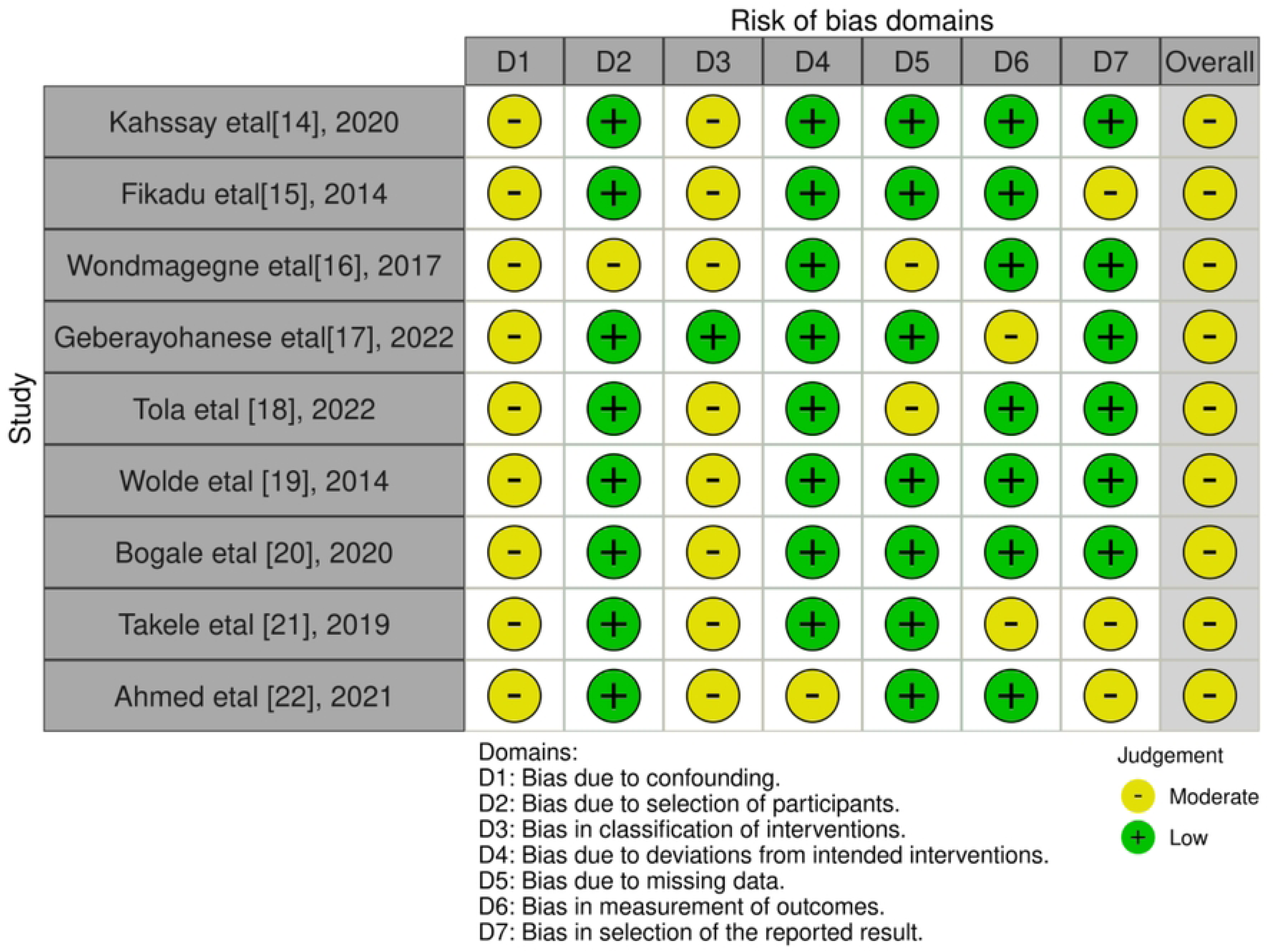
Risk of bias assessment using the ROBINS I tool for impact of breastfeeding on stunting : A systematic review and meta-analysis of observational studies in Ethiopia, 2000-2025.

### Results of synthesis

The random effect model meta-analysis’s overall pooled effect measure revealed that the lack of breastfeeding intervention increased risk of of stunting by [OR= 1.54 95%CI (0.94-2.13), P=<0.001]. Heterogeneity between studies was evaluated using the Higgins I^2^ statistical test, which indicated that [I^2^ = 86%] and the Cochran’s Q statistical test, which showed that [Q=34.44, df (degree of freedom) = 8 and P = < 0.001], which confirmed indicative of heterogeneity(Figure 3).

**Fig 3.**
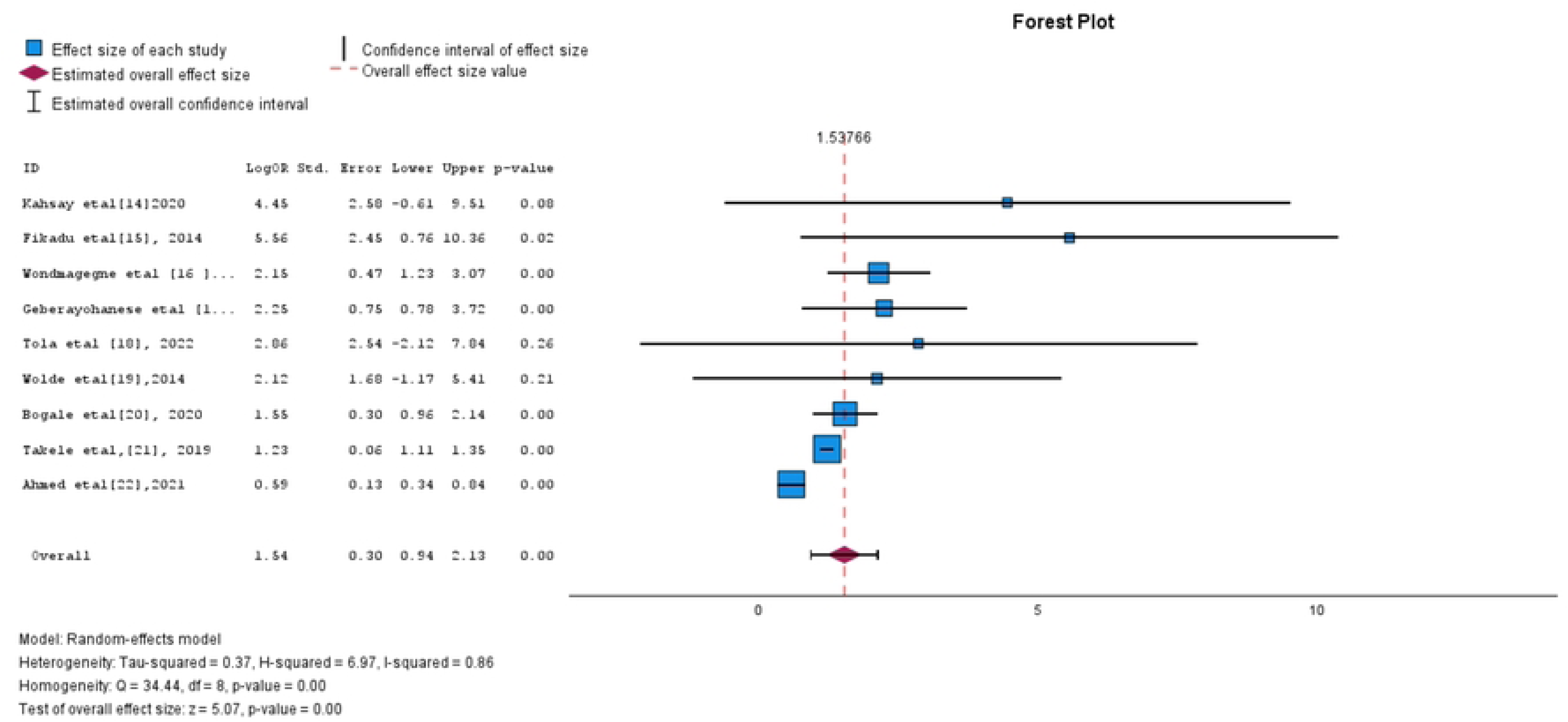
Forest plot of the impact of breastfeeding on stunting : A systematic review and meta-analysis of observational studies in Ethiopia, 2000-2025.

### Subgroup analysis

To determine the cause of the heterogeneity in the included studies, subgroup analysis was performed. Subgroup analysis revealed a statistically significant correlation between stunting and breastfeeding in multistage cluster and simple random sampling method. Research from the Southern area, Sidama, and Afar showed no statistical significance. A statistically significant correlation was discovered in both cross-sectional and case control study designs. Additionally, every sample size category displayed statistically significant results. However; study location such as Afar, Amhara, Oromiya, Sidama and Southern region, case control study design, sample size <1000 and 1000-5000; their effect measure were out of the confidence interval of the pooled effect measure. Hence, the source of the heterogeneity for the studies included in the review were study location, study design and sample size(Table 3).

**Table 3.** A leave - one - study - out sensitivity analysis on impact of breastfeeding on stunting : A systematic review and meta - analysis of observational studies in Ethiopia, 2000-2025.

| N - Excluded study | Effect | SE | P value | 95%CI<br>Lower | 95% CI<br>Upper | I <sup>2</sup> | Q test( P value) |
| --- | --- | --- | --- | --- | --- | --- | --- |
| Kahssay etal [14], 2020 | 1.43 | 0.28 | <0.001 | 0.77 | 2.09 | 82% | <0.001 |
| Fikadu etal [15], 2014 | 1.46 | 0.29 | <0.001 | 0.89 | 2.03 | 86% | <0.001 |
| Wondmagegne etal [16], 2017 | 1.39 | 0.31 | <0.001 | 0.78 | 1.99 | 84% | <0.001 |
| Geberayohanese etal [17], 2022 | 1.45 | 0.31 | <0.001 | 0.84 | 2.16 | 84% | <0.001 |
| Tola etal [18], 2022 | 1.46 | 0.30 | <0.001 | 0.75 | 2.16 | 83% | <0.001 |
| Wolde etal [19], 2014 | 1.46 | 0.30 | <0.001 | 0.77 | 2.18 | 83% | <0.001 |
| Bogale etal [20], 2020 | 1.54 | 0.36 | <0.001 | 0.69 | 2.39 | 86% | <0.001 |
| Takele etal [21], 2019 | 1.66 | 0.37 | <0.001 | 0.80 | 2.53 | 65% | <0.001 |
| Ahmed etal [22], 2021 | 1.27 | 0.08 | <0.001 | 1.10 | 1.45 | 0% | 0.11 |

N= total number of study

### Sensitivity analysis

For further investigation of the influence of each study in the meta-analysis of impact of breastfeeding on stunting; we performed a leave one study out sensitivity analysis. Our sensitivity analysis result showed that, the point estimated effect size of obtained when each study left out from meta-analysis was inside the confidence interval of the pooled effect size of the overall study and showed statistically significant association of breastfeeding and stunting. However; when excluded study (22) it showed not statistically significant association. All studies were showed heterogeneous, except study(22), which showed absence of heterogeneity [I^2^ = 0%]. The original pooled effect size was [OR=1.54, 95%CI (0.94-2.13)]. However, the leave out one study meta-analysis effect measure range showed between [OR= 1.27, (95%CI (1.10-1.45)] and [OR= 1.66, 95%CI(0.80-2.53)]. Therefore, study(22) were influential and affect the meta analysis significantly and the result of the leave out one study meta-analysis was not robust (Table 4).

### Publication biases

Begg’s funnel plot showed that it was asymmetrical and Egger’s regression test effect measure was [OR= 1.38, 95%CI (0.17-2.59) P= 0.03], provided then significant publication bias (Fig. 3). We applied the trim and fill method to adjust and enhance symmetry. Indicated the adjusted pooled effect determined by slop of egger’s regression test, using estimator of Restricted Maximum Likelihood (REML), revealed a pooled effect of [OR= 1.31, 95%CI (0.79-1.82) P= <0.001] (Figure 4).

**Fig 4.**
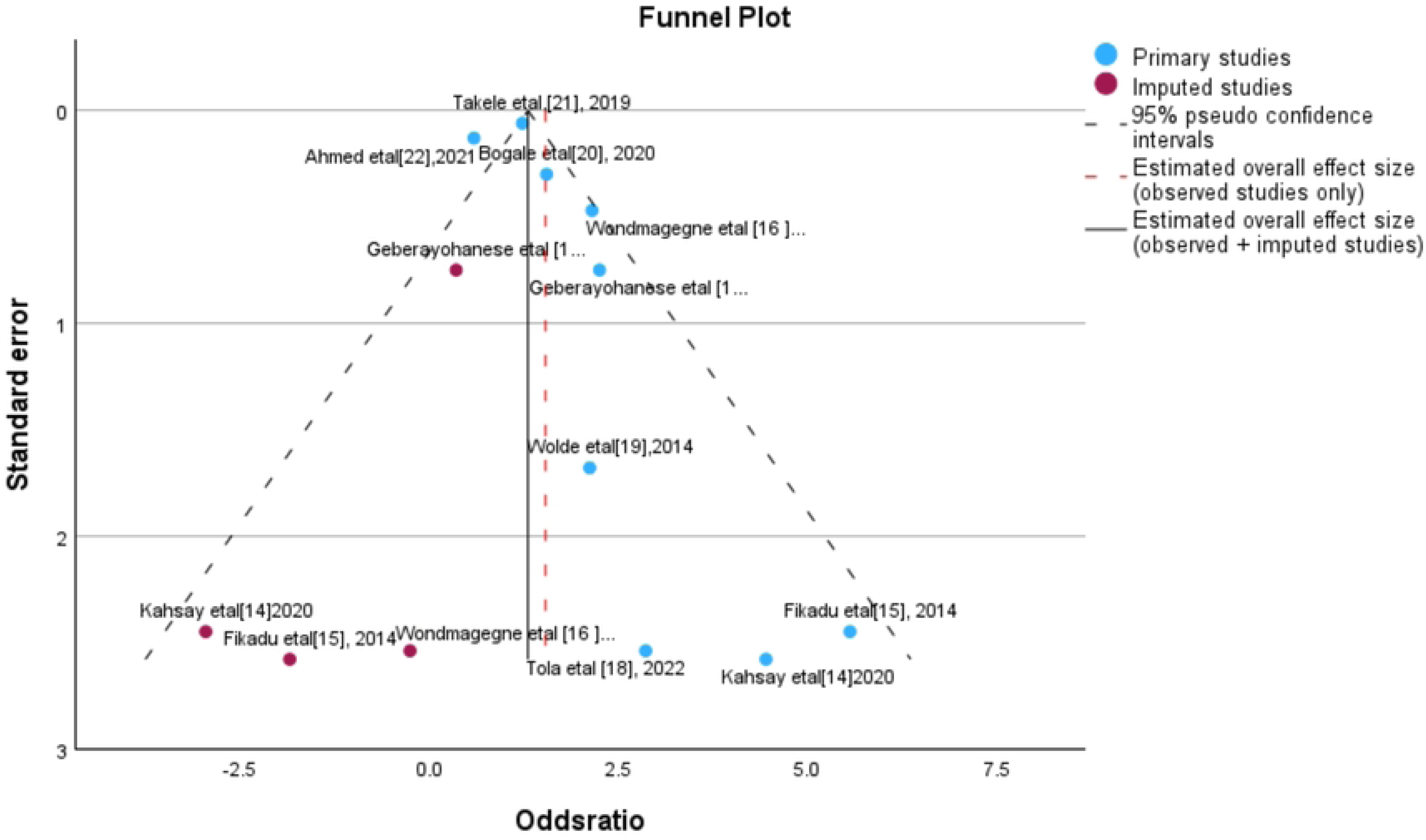
Begg’s funnel plot of the impact of breastfeeding on stunting : A systematic review and meta-analysis of observational studies in Ethiopia, 2000-2025.

### Certainty of evidence

We utilized GRADE technique (29) and GRADEpro software (30) to analyze certainty evidence of the study influence of breastfeeding on stunting. Generally, the evidence assessment’s overall conclusion indicated that its quality was moderate.

## Discussion

Nine studies were found and evaluated by this systematic review and meta-analysis using database search conducted between January, 2000 and July, 2025; however, the included studies were published between 2014 and 2022. Upon examining the individual study designs, we found that six of the studies reported a statistically significant association between breastfeeding and stunting (15–17, 20–22), while three studies reported no statistically significant association (14, 18, 19). Each study was subjected to a risk of bias assessment using the ROBINS I tool (26) with seven domains of bias, and the overall risk of bias assessment for the studies were rated as moderate.

In Ethiopia, it was found that not breastfeeding increased the incidence of stunting with a pooled impact of [OR = 1.54, 95%CI(0.94-2.13), P=<0.001]. This result was also found to be in line with meta-analyses and comprehensive reviews carried out in various nations (43-49). Additionally, clinical research showed that not providing breastfeeding reduce key nutrient that support child’s optimal growth and immune boosting proteins component helps prevent a child from various disease, and consequently increases the risk of stunting.

The Cochrane’s Q statistical test revealed heterogeneity among the studies, with [Q=34.44, df =8, P = <0.001] and the Higgins I^2^ statistical test revealed that [I^2^= 86%], which was verified to be a sign of significant heterogeneity. As a result, the random effect model was the meta analysis model utilized to analyze the pooled effect size.

Subgroup analysis revealed a statistically significant correlation in both cross-sectional and case control study designs, all sample size categories, and multistage cluster and simple random sampling method. However, study locations such as Afar, Amhara, Oromiya, Sidama, and Southern region, as well as case control study designs and sample sizes <1000, and 1000-5000, had effect measures that were outside the range of the confidence interval of the pooled effect measure. Therefore, the source of the heterogeneity for the studies included in the review was the combination of study location, study design, and sample size.

The estimated effect size derived from excluding each study from the meta-analysis fell within the confidence interval of the whole study’s pooled effect size, according to the results of the sensitivity analysis. All of the sensitivity analysis, with the exception of study (22) were statistically significant. Furthermore, all the leave out one study analysis showed heterogeneity, with the exception of study (22), which reported no heterogeneity (I^2^ = 0%). The pooled effect measure ranges of the sensitivity analysis between [OR= 1.27, 95%CI (1.10-1.45)] and [OR= 1.66, 95%CI (0.80-2.53)]. The findings of the leave-out one study meta analysis were therefore not robust, and study (22) was an influential study.

When it came to publication bias, Begg’s funnel plot was reported as asymmetrical (Fig. 3). Egger’s regression test effect measure was given [OR= 1.38, 95%CI (0.17-2.59) P= 0.03] and revealed presence of publication bias. As a result, we used the trim and fill technique to improve and modify symmetry using the Restricted Maximum Likelihood (REML) estimator, the adjusted pooled effect as assessed by the slop of Egger’s regression test showed a pooled effect of [OR= 1.31, 95%CI (0.79-1.82) P= <0.001]. The GRADE approach was used to evaluate the studies’ evidence quality, which revealed that it was of moderate quality.

### Strength and limitation of the study

One of the strength of this study was it adhere with PRISMA guideline. Secondly, the this systematic review and meta analysis employed ROBINS-I tool and GRADE approach to assess the risk of bias and certainty of evidence of the studies included respectively which helps to assess quality of the studies. On other side, the limitation of the evidence included in this systematic review and meta analysis was the studies included were heterogeneous due to sample size, study design, and study location difference. Secondly, the pooled effect size may be impacted by the moderate quality of the study’s evidence. Third, the review also noted the existence of publication bias, which could have an impact on the findings of the meta-analysis and systematic review.

## Conclusion and recommendation

According to the results of the meta-analysis, not breastfeeding is a risk factor for a higher incidence of stunting in Ethiopia. In order to encourage breastfeeding and prevent the high prevalence of stunting in Ethiopia, health officials should incorporate breastfeeding policies into their nutrition programs.

## Data Availability

All data produced in the present work are contained in the manuscript

## Acknowledgment

The authors would like to thank all the authors and publishers of original studies.

## Notes

### Competing Interest Statement

The authors have declared no competing interest.

### Clinical Protocols

https://www.crd.york.ac.uk/PROSPERO/view/CRD420251108544)

### Funding Statement

This study did not receive any funding

